# Vaccine effectiveness against SARS-CoV-2 infection with the Omicron or Delta variants following a two-dose or booster BNT162b2 or mRNA-1273 vaccination series: A Danish cohort study

**DOI:** 10.1101/2021.12.20.21267966

**Authors:** Christian Holm Hansen, Astrid Blicher Schelde, Ida Rask Moustsen-Helm, Hanne-Dorthe Emborg, Tyra Grove Krause, Kåre Mølbak, Palle Valentiner-Branth, on behalf of the Infectious Disease Preparedness Group at Statens Serum Institut

## Abstract

In this brief communication we are showing original research results with early estimates from Danish nationwide databases of vaccine effectiveness (VE) against the novel SARS-CoV-2 Omicron variant (B.1.1.529) up to five months after a primary vaccination series with the BNT162b2 or mRNA-1273 vaccines.

Our study provides evidence of protection against infection with the Omicron variant after completion of a primary vaccination series with the BNT162b2 or mRNA-1273 vaccines; in particular, we found a VE against the Omicron variant of 55.2% (95% confidence interval (CI): 23.5 to 73.7%) and 36.7% (95% CI: -69.9 to 76.4%) for the BNT162b2 and mRNA-1273 vaccines, respectively, in the first month after primary vaccination. However, the VE is significantly lower than that against Delta infection and declines rapidly over just a few months. The VE is re-established upon revaccination with the BNT162b2 vaccine (54.6%, 95% CI: 30.4 to 70.4%).

## INTRODUCTION

On 26 November, 2021, the severe acute respiratory syndrome coronavirus 2 (SARS-CoV-2) variant B.1.1.529, named Omicron, was classified as a variant of concern by the WHO and is currently spreading rapidly across the globe including in Denmark.^1,2^ Here, we estimate COVID-19 vaccine protection against infection with Omicron or Delta up to five months after primary vaccination using Danish nationwide data.

## METHODS

Complete residency, COVID-19 PCR test and vaccination data were extracted from Danish nationwide registries. Nearly all PCR-confirmed COVID-19 cases identified in the country during the study period (November 20 to December 12, 2021) were investigated for Omicron by whole-genome sequencing or a novel variant specific PCR test targeting the 452L mutation. Cases not identified as Omicron were assumed to be Delta.

Vaccine effectiveness (VE) was estimated in a time-to-event analysis of Danish residents ≥12 years comparing the rate of infections in unvaccinated and vaccinated individuals with a two-dose BNT162b2 or mRNA-1273 vaccination series. Unvaccinated individuals were followed up from November 20th. Vaccinated individuals contributing to the estimate for the first period (1-30 days after full protection) were followed up from November 20th or, if later, 14 days after receiving their second dose, and were observed until December 12th or, if earlier, 30 days after full protection (44 days after the second dose). Vaccinated individuals contributing to the estimates for the three subsequent periods were similarly observed respectively from day 31-60, 61-90 and 91-150 after full protection. Follow-up was censored at the earliest of December 12th, a positive SARS-CoV-2 PCR test, emigration, death, vaccination for unvaccinated or revaccination for vaccinated individuals. Previously SARS-CoV-2 PCR-positive individuals were excluded.

VE was calculated as 1-HR with HR (hazard ratio) estimated in a Cox regression model adjusted for age, sex and geographical region, and using calendar time as the underlying time scale.

## RESULTS

By December 12, 2021, there were 5,767 identified Omicron cases in Denmark with a median age of 28 years (93% <60 years). Among those who had most recently completed primary vaccination, VE against Omicron was 55.2% (95% confidence interval: 23.5 to 73.7%) and 36.7% (−69.9 to 76.4%) for the BNT162b2 and mRNA-1273 vaccines, respectively, but with evidence of rapid waning over the course of five months. By comparison, VE against Delta was significantly higher and better preserved over the same period (see Figure and Table).

**Figure.**
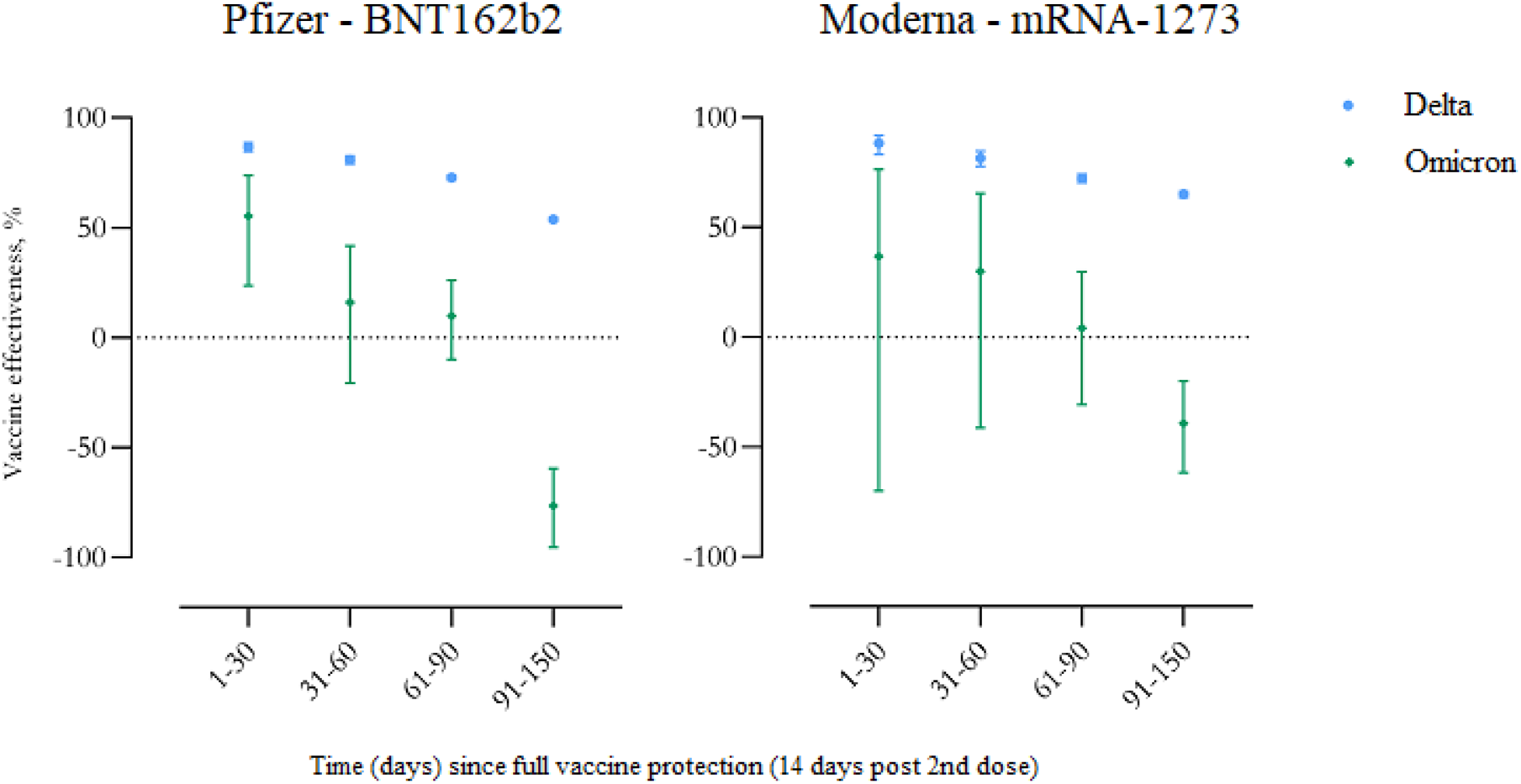
Vaccine effectiveness against SARS-CoV-2 infection with the Delta and Omicron variants, shown separately for the BNT162b2 and mRNA-1273 vaccines. Vertical bars indicate 95% confidence intervals.

**Table.**
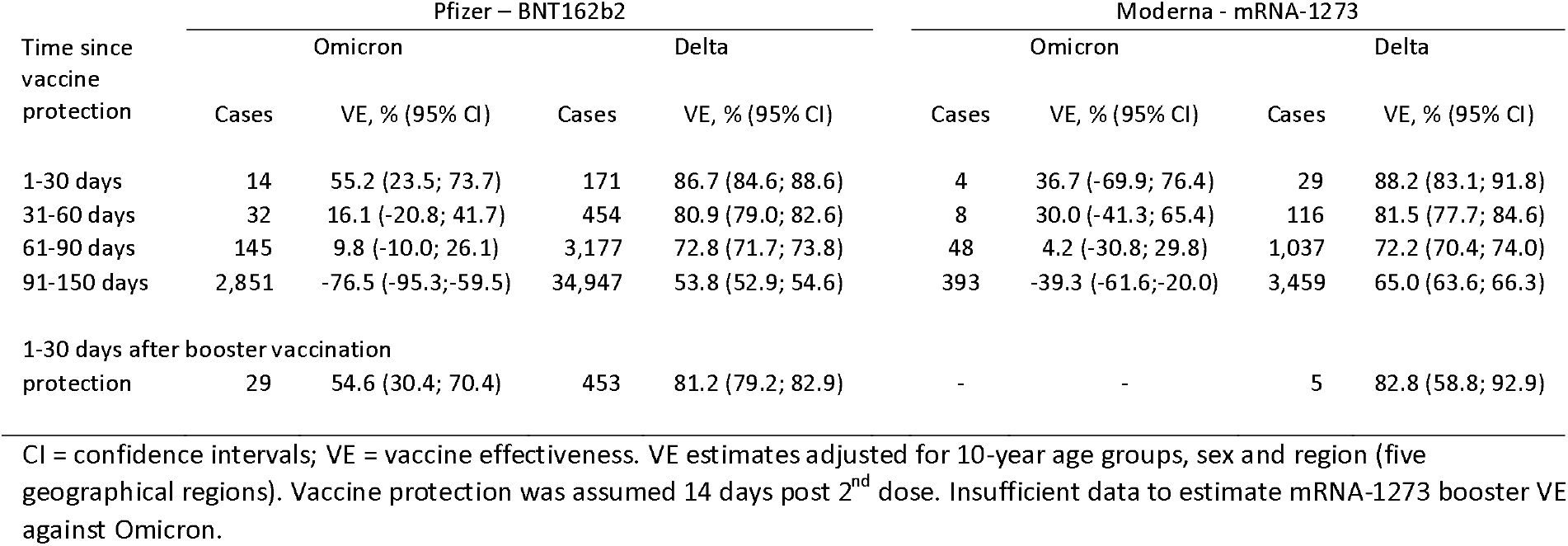
Estimated vaccine effectiveness for BNT162b2 and mRNA-1273 against infection with the SARS-CoV-2 Omicron and Delta variants during November 20 – December 12, 2021, Denmark.

VE among those who had received a booster dose 14 to 44 days earlier was 54.6% (30.4 to 70.4%) using those with only primary vaccination as comparison (analysis restricted to 60+ year-olds).

## DISCUSSION

VE against Omicron was 55.2% initially following primary BNT162b2 vaccination, but waned quickly thereafter. Although estimated with less precision, VE against Omicron after primary mRNA-1273 vaccination similarly indicated a rapid decline in protection. By comparison, both vaccines showed higher, longer-lasting protection against Delta. The negative estimates in the final period arguably suggest different behaviour and/or exposure patterns in the vaccinated and unvaccinated cohorts causing underestimation of the VE. This was likely the result of Omicron spreading rapidly initially through single (super-spreading) events causing many infections among young, vaccinated individuals.

A recent study from England (in preprint) found higher effectiveness against symptomatic Omicron initially after BNT162b2 vaccination followed by a rapid decline in protection, and that VE increased to 75.5% (56.1 to 86.3%) two weeks after booster vaccination using unvaccinated individuals as comparison.

Our study contributes to emerging evidence that BNT162b2 or mRNA-1273 primary vaccine protection against Omicron decreases quickly over time with booster vaccination offering a significant increase in protection. In light of the exponential rise in Omicron cases, these findings highlight the need for massive rollout of vaccinations and booster vaccinations.

## Data Availability

De-identified data are available for access to members of the scientific and medical community for non-commercial use only upon reasonable request to the authors

